# MHC Haplotyping of SARS-CoV-2 patients: HLA subtypes are not associated with the presence and severity of Covid-19 in the Israeli population

**DOI:** 10.1101/2020.12.23.20248148

**Authors:** Shay Ben Shachar, Noam Barda, Sigal Manor, Sapir Israeli, Noa Dagan, Adam Haber, Shai Carmi, Ran Balicer, Bracha Zisser, Yoram Louzoun

## Abstract

HLA haplotypes were found to be associated with increased risk for viral infections or disease severity in various diseases, including SARS. Several genetic variants are associated with Covid-19 severity. However, no clear association between HLA and Covid-19 incidence or severity has been reported. We conducted a large scale HLA analysis of Israeli individuals who tested positive for SARS-CoV-2 infection by PCR. Overall, 72,912 individuals with known HLA haplotypes were included in the study, of whom 6,413 (8.8%) were found to have SARS-CoV-2 by PCR. a Total of 20,937 subjects were of Ashkenazi origin (at least 2/4 grandparents). One hundred eighty-one patients (2.8% of the infected) were hospitalized due to the disease. None of the 66 most common HLA loci (within the five HLA subgroups; A, B, C, DQB1, DRB1) was found to be associated with SARS-CoV-2 infection or hospitalization. Similarly, no association was detected in the Ashkenazi Jewish subset. Moreover, no association was found between heterozygosity in any of the HLA loci and either infection or hospitalization.

We conclude that HLA haplotypes are not a major risk/protecting factor among the Israeli population for SARS-CoV-2 infection or severity.

## MAIN TEXT

The Major Histocompatibility Complex (MHC) molecules are cell surface protein complexes encoded in the Human Leukocyte Antigens (HLA) locus. The HLA locus is highly polymorphic with thousands of different alleles and millions of haplotypes reported^1,2^. HLAs are associated with several infectious diseases. Previous disease association studies showed that some HLA haplotypes are highly correlated with multiple viral infections^3,4,5^ and have been reported to confer differential susceptibility to infection and the severity of the resulting disease^6^. In particular, HLA class I alleles are key players in the immune defense against viruses as they initiate the activity of T cells against the invading intracellular pathogens ^7,8^. Although presentation of viral antigens relies classically on MHC class I molecules, MHC class II genes have also been associated with the outcome of many viral infections^9,10,11^. Studies in a range of species^12,13,14,15^, including humans^16,17^, imply a heterozygous selection mechanism operating on the HLA loci as an explanation for the extensive variability of the MHC molecules^18,19^. However, recent results on human populations suggest that such an advantage may be limited^20^.

In the context of Covid-19, specific HLAs have been associated with coronaviruses in the past, including SARS coronavirus infections^21,22^. Several efforts were made to identify HLA-related susceptibility to SARS-CoV-1 after the first SARS epidemic in East Asia^23,24,25,26^ and the MERS-CoV outbreak in 2014 in Saudi Arabia^27^. Most of these case studies provided weak or conflicting results and required further validation due to the relatively small sample size. An association between the geographic distributions of Covid-19 and HLA alleles has been suggested^28^. Similarly, an association between severe disease outcome and specific HLAs was suggested by another small-scale whole-genome sequencing study^29^. In silico model raised the possibility that HLA supertypes may have a role in the severity of the disease^30^. Moreover, a reduced expression of human leukocyte antigen class DR (HLA-DR) was detected in peripheral blood mononuclear cells of Covid-19 patients^31^. Nevertheless, genome-wide association study (GWAS) among 1980 Italian and Spanish patients with severe Covid-19 detected a few loci associated with that state but no association with the HLA loci at chromosome 6 or heterozygote advantage was found^32^. However another GWAS study analyzing 2244 critically-ill Covid-19 patients in the UK detected an association between HLA-G and HLA-DPB1 and Covid-19^33^. Two other large unpublished GWAS studies did not report a signal at the HLA locus on chromosome 6^34,35^. To the best of our knowledge there are no previous studies looking specifically at HLA subgroups among individuals with Covid-19.

HLA allele and haplotype frequencies differ between populations belonging to different ethnic groups^36,37^. For example, among Jewish populations, similarities of HLA alleles may be seen within ethnicities, allowing an estimation of an individual’s origin based on his HLA genotypes^38^. It has been estimated that decreased HLA variability may assist to detect signals relevant to a specific disease state. Indeed, specific HLAs were associated with different disorders in the Ashkenazi population, a large genetically isolated group^39,40,41^. Given the diversity of HLAs among populations, testing genetically isolated and genetically homogenous populations may provide an advantage in detecting the effects of specific genetic variants, in particular in Ashkenazi Jews.

Our study aimed to utilize the presence of a large-scale cohort having HLA genotypes and a high rate of Covid-19 to test the possible role of specific HLA alleles in SARS-CoV-2 infection and its severity. Using a data set of this magnitude also allowed us to analyze associations between Covid-19 infection and HLA zygosity. Understanding how variation in HLA may affect both susceptibility and severity of Covid-19 infection could help to identify and stratify individuals at higher risk for the disease and may support future vaccination strategies.

A case-control study with a test-negative design^42^ was used to evaluate the infection outcome, while a retrospective cohort study was used to evaluate the hospitalization outcome. In the case-control design, patients tested positive or negative for SARS-CoV-2 were evaluated for their HLA alleles. In the cohort design, patients who were diagnosed with COVID-19 were followed-up to see who were hospitalized.

The study was based on patient data taken from the data warehouse of Clalit Health Care (CHS), a large integrated payer-provider healthcare organization operating in Israel. CHS insures over 4.6 million Israelis (∼50%), which are a representative sample of the entire population.

HLA alleles were obtained from the Ezer Mizion Bone Marrow Donor Registry, which enlists the highest number of registered unrelated volunteer donors per capita in the world. The HLA resolution varies from serologic to DNA-based testing at low, intermediate, and allele resolution^43^. The initial analysis included HLA genotypes of 1,040,250 donors which represent all the populations composing the Ezer Mizion registry. High-resolution five-locus typing was imputed for all donors using GRIMM (GRaph IMputation and Matching for HLA Genotypes)^44^, which produces the most likely five-locus genotype consistent with the low-resolution typing and the self-defined ethnicity. For each subject in the dataset, we chose the unphased genotype with the highest probability. We also used the low resolution two-digit typing of each subject, as well as the homozygosity status of each allele.

The study included all patients tested for SARS-CoV-2 by PCR that were members of CHS at the date of testing and had HLA data available. Cases were those patients tested positive, while controls were patients tested negative. Covariates for adjustment included age, sex, number of children in the household, population sector (ultra-orthodox vs. general, which has had an important impact on the epidemic in Israel), socioeconomic status, and ethnic origin (Ashkenazi vs. other). These covariates were extracted as of the test date. The number of children in the household was calculated based on CHS’ demographic registry. The population sector was determined using the patient’s primary-care clinic address. Socioeconomic status was determined based on a patient’s home address. Patients were defined as Ashkenazi if they were Jewish and were born themselves or had at least one parent or 2 grandparents born in central or western Europe. A severe disease course was defined by hospitalization less than 2 weeks after the molecular diagnosis.

The association between each HLA allele and the outcome, adjusted for the covariates listed above, was tested separately for each allele using logistic regression. Homozygosity was tested similarly, as a binary predictor, separately for each locus. Bonferroni correction was applied to the p-values and confidence intervals. The results were similar when the Benjamini-Hochberg false discovery rate was controlled^45^. Zygosity was computed based on two-digit HLA alleles at the appropriate locus. The study was performed with the approval of the Institutional Research Community (IRB) #0080-20-COM2).

Overall, 72,912 individuals with HLA genotyping and at least one PCR test for Covid-19 were included in the study between March 1^st,^ 2020 and October 31th 2020. Of them, 8.8% (6,413) tested positive for SARS-CoV-2 by PCR. Demographic and socioeconomic data related to the population is available in Table 1.

**Table 1:**
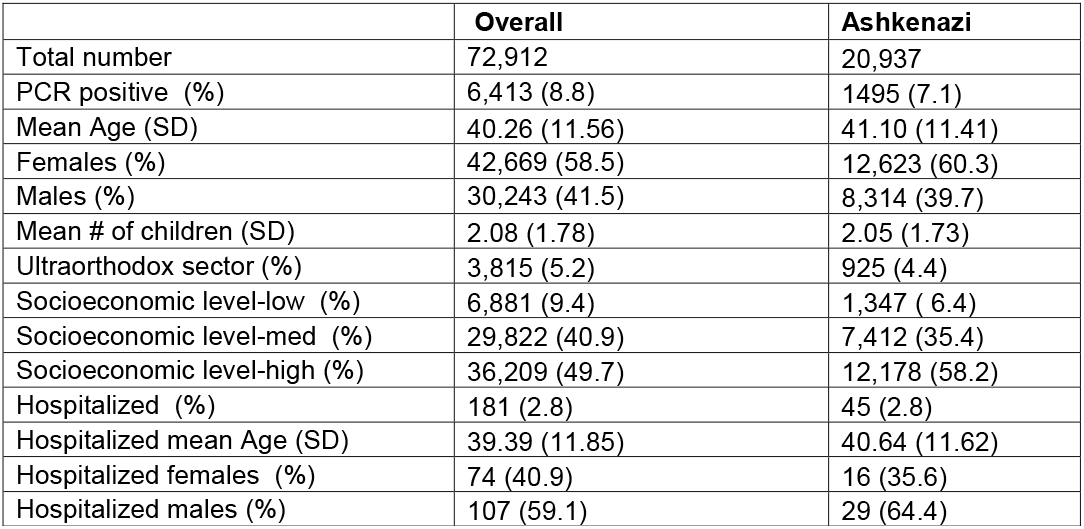
Demographic Characterization of study groups groups. Characterization of the individuals with HLA genotyping and at least one PCR test for Covid-19 that were included in the study.

|  | **Overall** | **Ashkenazi** |
| --- | --- | --- |
| Total number | 72,912 | 20,937 |
| PCR positive (%) | 6,413 (8.8) | 1495 (7.1) |
| Mean Age (SD) | 40.26 (11.56) | 41.10 (11.41) |
| Females (%) | 42,669 (58.5) | 12,623 (60.3) |
| Males (%) | 30,243 (41.5) | 8,314 (39.7) |
| Mean # of children (SD) | 2.08 (1.78) | 2.05 (1.73) |
| Ultraorthodox sector (%) | 3,815 (5.2) | 925 (4.4) |
| Socioeconomic level-low (%) | 6,881 (9.4) | 1,347 ( 6.4) |
| Socioeconomic level-med (%) | 29,822 (40.9) | 7,412 (35.4) |
| Socioeconomic level-high (%) | 36,209 (49.7) | 12,178 (58.2) |
| Hospitalized (%) | 181 (2.8) | 45 (2.8) |
| Hospitalized mean Age (SD) | 39.39 (11.85) | 40.64 (11.62) |
| Hospitalized females (%) | 74 (40.9) | 16 (35.6) |
| Hospitalized males (%) | 107 (59.1) | 29 (64.4) |

All HLA alleles were grouped by two-digit representations to minimize the number of tests, and only alleles with a frequency of at least 1 % were used. A total of 16 HLA-A, 20 HLA-B, 13 HLA-C, 5 HLA-DQB1, and 12 HLA-DRB1 (Table 2) met the criteria for analysis. No significant differences in HLA-allele frequency were detected between individuals with negative and positive SARS-CoV-2 PCR (Figure 1 Left panel).

**Table 2:** HLA-subgroups (2-digit) analyzed. List of two-digit HLA alleles with a population frequency >1%, analyzed in the study

|  | **HLA-A** | **HLA-B** | **HLA-C** | **HLA-DQB1** | **HLA-DRB1** |
| --- | --- | --- | --- | --- | --- |
|  | A_1 | B_7 | C_1 | DQ_2 | DR_1 |
|  | A_2 | B_8 | C_2 | DQ_3 | DR_3 |
|  | A_3 | B_13 | C_3 | DQ_4 | DR_4 |
|  | A_11 | B_14 | C_4 | DQ_5 | DR_7 |
|  | A_23 | B_15 | C_5 | DQ_6 | DR_8 |
|  | A_24 | B_18 | C_6 |  | DR_10 |
|  | A_25 | B_27 | C_7 |  | DR_11 |
|  | A_26 | B_35 | C_8 |  | DR_12 |
|  | A_29 | B_38 | C_12 |  | DR_13 |
|  | A_30 | B_40 | C_14 |  | DR_14 |
|  | A_31 | B_41 | C_15 |  | DR_15 |
|  | A_32 | B_44 | C_16 |  | DR_16 |
|  | A_33 | B_49 | C_17 |  |  |
|  | A_66 | B_50 |  |  |  |
|  | A_68 | B_51 |  |  |  |
|  | A_69 | B_52 |  |  |  |
|  |  | B_53 |  |  |  |
|  |  | B_55 |  |  |  |
|  |  | B_57 |  |  |  |
|  |  | B_58 |  |  |  |

**Figure 1:**
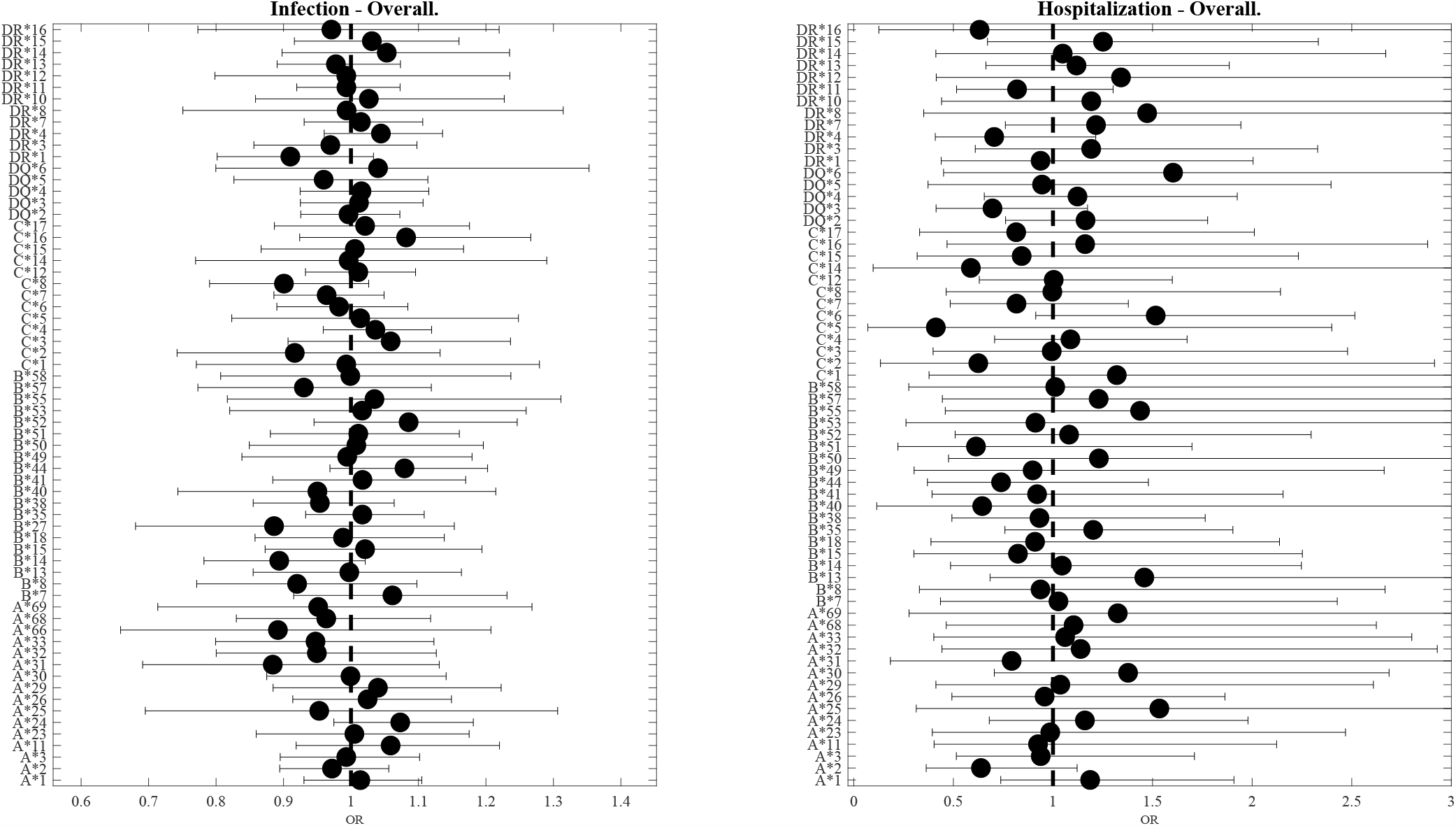
Odds Ratio (OR) as a function of HLA allele. In all figures, the dot represents the odds ratio, and the interval represents the 95% confidence intervals (Bonferroni correction). The left plot is for SARS-CoV-2 infection and the right plot is for hospitalization. We trim error bars at 3 for better visualization. The analysis presented here is for the full population. One can see that no HLA allele is significantly associated.

We further tested whether HLA genotypes may be associated with severe Covid-19 infection. Of the 6,413 infected individuals, 181 (2.6%) were hospitalized within 2 weeks. Again, no significant HLA genotypes were found to be associated with hospitalization among individuals positive for SARS-CoV-2 (Figure 1 Right panel).

Given the association between HLA and ethnic origin and the fact that the Ashkenazi Jewish population is genetically isolated^35^, we tested for SARS-CoV-2 presence and severity within the Ashkenazi population. This analysis included 20,937 Ashkenazi donors, of which 1,495 were positive for SARS-CoV-2, and 45 were hospitalized. No significant association was observed between either SARS-CoV-2 positive frequency (Figure 2 Left panel) or Covid-19 severity (Figure 2 Right panel) and any HLA subgroup in the Ashkenazi population (Figure 2).

**Figure 2:**
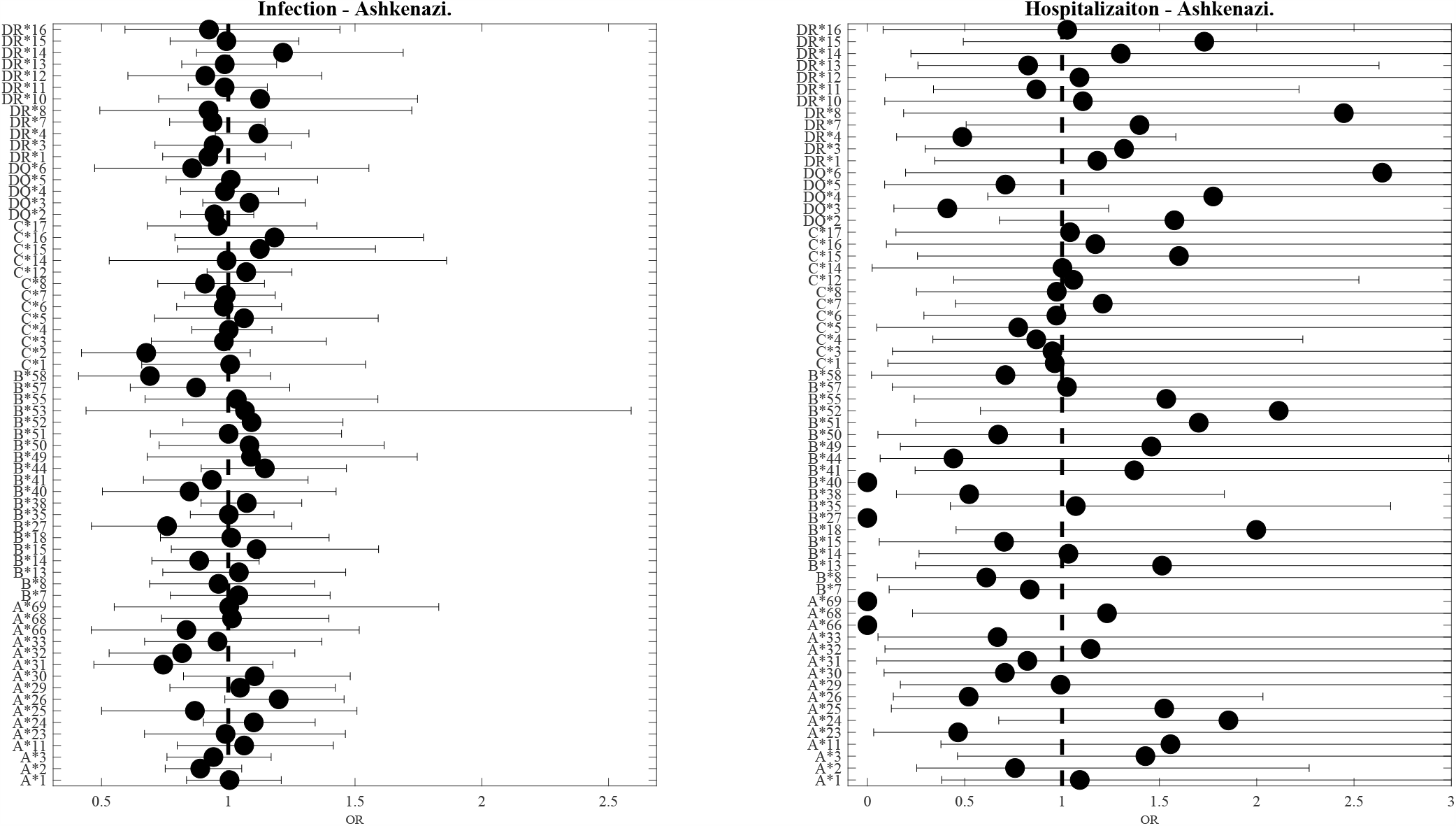
Odds Ratio (OR) as a function of HLA allele in the Ashkenazi population. In all figures, the dot represents the odds ratio, and the interval represents the 95% confidence intervals (Bonferroni correction). The left plot is for SARS-CoV-2 infection and the right plot is for hospitalization. We trim error bars at 3 for better visualization. The analysis presented here is for the Ashkenazi population. One can see that no HLA allele is significantly associated

To investigate whether HLA heterozygosity is associated with the outcome of SARS-CoV-2 infection we further tested whether an increased probability of SARS-CoV-2 infection or severe disease course are associated with a higher homozygous rate of each of the 5 HLA loci subgroup. No such association was detected either in the general population or in the Ashkenazi population (Figure 3).

**Figure 3:**
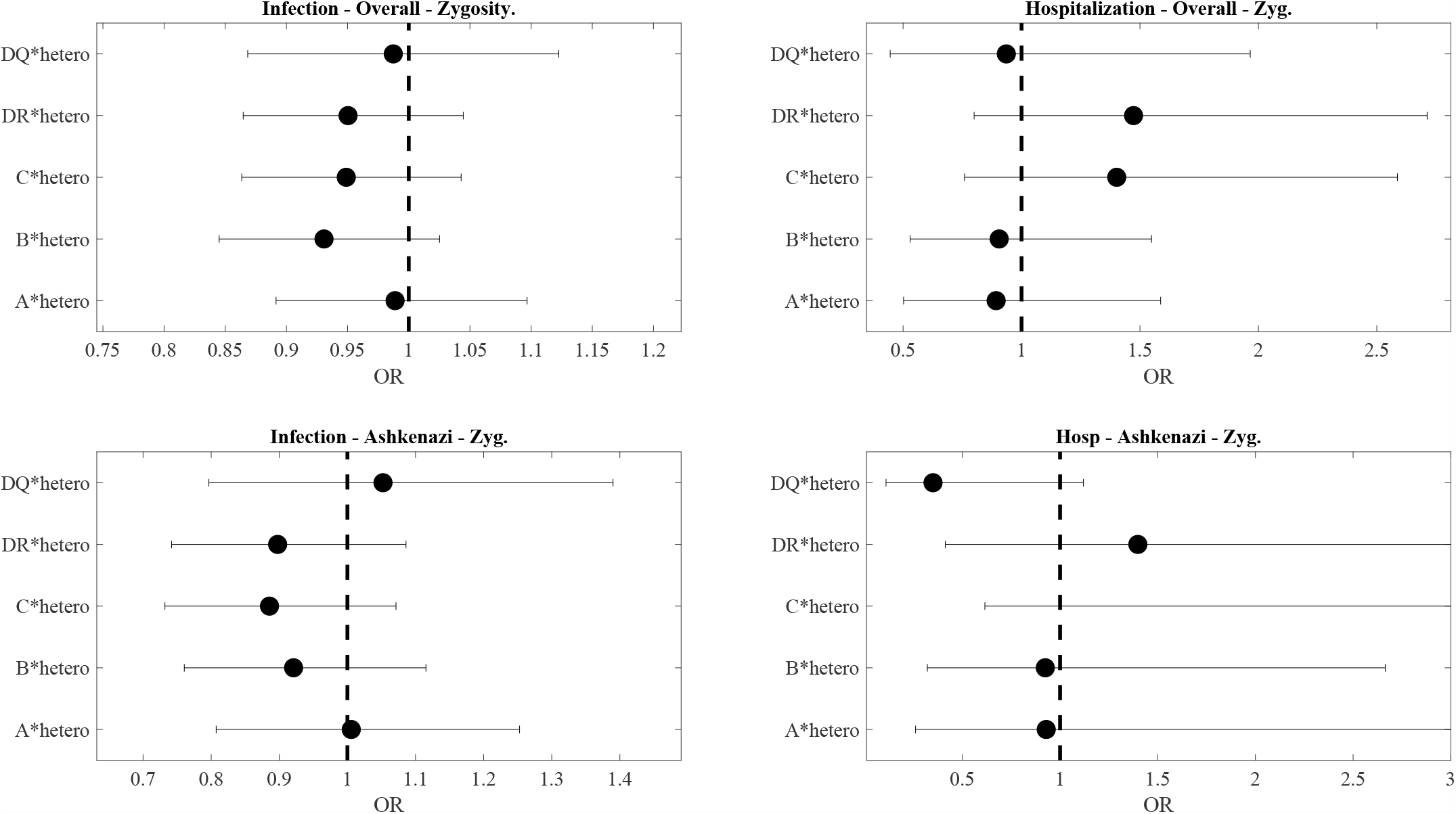
Effect of homozygosity on either infection or hospitalization in the entire studied population or the Ashkenazi population. Again no significant association was found. Homozygosity at a given locus is defined as having the same low-resolution allele in this locus.

The Covid-19 pandemic is associated with heavy medical, economic, and social costs. A few factors were associated with decreased or increased risks of SARS-CoV-2 infection, including smoking^46^ and gender (male)^47^. Genetic factors were also found to be associated with Covid-19 disease severity ^48^, including a genomic segment of around 50 kilobases in size that is inherited from Neanderthals^49^. Similarly, a meta-analysis of GWAS studies of patients from Spain and Italy with severe Covid-19, which was defined as a hospitalization with respiratory failure, detected three significant loci with odd ratios of 1.32-1.77 per loci^32^. Given the important role of HLA in several viral infections, the above study analyzed the extended HLA region (chromosome 6, 25 through 34 Mb) but did not find significant SNP association signals at the HLA complex.

Our study, performed on a large scale dataset, did not detect any specific HLA subgroups associated with increased or decreased risk for SARS-CoV-2 infection, or a severe disease course causing hospitalization. We also did not detect any associations of heterozygote advantage with Covid-19 when we analyzed the entire study population or when analyzing a sub-population. In contrast with GWAS studies, in which a very large number of loci are compared and thus increasing the multiple testing burden, we incorporated HLA alleles with a prevalence of >1% in the Israeli population. Ths approach enabled a large-scale analysis not of a few HLA subtypes but of f 66 variants.

Two main caveats should be considered. First, the imputation of HLA alleles may induce errors. However, the precision of the imputation was previously established^50^. Moreover, no association was found with the A alleles that were typed in the vast majority of subjects. Finally, the analysis was performed at the two-digit level, which is practically unaffected by imputation. Another possible limitation is the age distribution. Our study included only a small proportion of hospitalized cases 181/6,413 (2.6%). This may be related to the average young age of individuals for whom we have HLA data. However, this should not affect the results related to SARS-CoV-2 infection. While hospitalization alone may not be an indicator of disease severity, we expect a significant enrichment of severe cases in hospitalized patients.

The lack of HLA association reported here and in previous smaller studies, and the absence of advantage for heterozygote may suggest that heterozygote advantage in HLA may be more limited in general than often considered. This is in agreement with more recent claims that other mechanisms beyond heterozygote advantage or pathogen-driven balancing selection may be the source of the large HLA polymorphism^2,20^.

We did not detect an HLA association with the initial phases of infection and hospitalization. As in other diseases, the initial response to the disease in the first few days is mainly mediated by the innate immune response, limiting the involvement of HLA and T Cells. HLA may still be involved in the later phases of the disease progression, such as long term sequels, or complications following hospitalization. A long-term follow-up will be needed to test for those.

## Data Availability

Data will be available upon request

## Data. Availability

The frequency of each allele in each dataset can be obtained from the authors upon demand.

